# Potential Utility of C-reactive Protein for Tuberculosis Risk Stratification among Patients with Non-Meningitic Symptoms at HIV Diagnosis in Low- and Middle-Income Countries

**DOI:** 10.1101/2023.12.19.23300232

**Authors:** Kathryn Dupnik, Vanessa R. Rivera, Nancy Dorvil, Hanane Akbarnejad, Yipeng Gao, Jingyi Liu, Alexandra Apollon, Emelyne Dumond, Cynthia Riviere, Patrice Severe, Kerlyne Lavoile, Maria Alejandra Duran Mendicuti, Samuel Pierre, Vanessa Rouzier, Kathleen F. Walsh, Anthony L. Byrne, Patrice Joseph, Pierre-Yves Cremieux, Jean William Pape, Serena P. Koenig

**Affiliations:** Weill Cornell Medicine, New York, New York, USA; Haitian Group for the Study of Kaposi’s Sarcoma and Opportunistic Infections (GHESKIO), Port-au-Prince, Haiti; The Analysis Group, Boston, Massachusetts, USA; Brigham and Women’s Hospital, Harvard Medical School, Boston, Massachusetts, USA; St. Vincent’s Hospital and Clinical School, University of New South Wales, Darlinghurst, New South Wales, Australia

**Author notes:** Corresponding Author Serena P. Koenig, Brigham and Women’s Hospital 75 Francis Street, Boston, MA 02115, USA.

**Keywords:** HIV, tuberculosis, C-reactive protein

## Abstract

**Article Summary:** We assessed the association between C-reactive protein (CRP) and *Mycobacterium tuberculosis* (TB) diagnosis in symptomatic patients at HIV diagnosis. We found that CRP concentrations can improve tuberculosis risk stratification, facilitating decision making about whether (specific) tuberculosis testing is indicated before antiretroviral therapy initiation.

**Background:** The World Health Organization recommends initiating same-day ART while tuberculosis testing is underway for patients with non-meningitic symptoms at HIV diagnosis, though safety data are limited. C-reactive protein (CRP) testing may improve tuberculosis risk stratification in this population.

**Methods:** In this baseline analysis of 498 adults (>18 years) with tuberculosis symptoms at HIV diagnosis who were enrolled in a trial of rapid ART initiation in Haiti, we describe test characteristics of varying CRP thresholds in the diagnosis of TB. We also assessed predictors of high CRP (≥3 mg/dL) using generalized linear models.

**Results:** Eighty-seven (17.5%) patients were diagnosed with baseline TB. The median CRP was 33.0 mg/L (IQR: 5.1, 85.5) in those with TB, and 2.6 mg/L (IQR: 0.8, 11.7) in those without TB. As the CRP threshold increased from ≥1 mg/L to ≥10 mg/L, the positive predictive value for TB increased from 22.4% to 35.4%, and negative predictive value decreased from 96.9% to 92.3%. With CRP thresholds varying from <1 to <10 mg/L, a range from 25.5% to 64.9% of the cohort would have been eligible for same-day ART, and 0.8% to 5.0% would have untreated TB at ART initiation.

**Conclusions:** CRP concentrations can be used to improve TB risk stratification, facilitating same-day decisions about ART initiation. Depending on the CRP threshold, one-quarter to two-thirds of patients could be eligible for same-day ART, with a reduction of 3-fold to 20-fold in the proportion with untreated TB, compared with a strategy of same-day ART while awaiting TB test results.

## INTRODUCTION

Global guidelines recommend antiretroviral therapy (ART) initiation on the same day of HIV diagnosis for patients who are ready to start treatment.^1–4^ These guidelines have been implemented in response to several clinical trials and cohort studies, which have demonstrated improved outcomes and faster time to viral suppression with this approach.^5–9^ However, a substantial proportion of patients in high burden settings present with symptoms of tuberculosis (TB) at HIV diagnosis.^10,11^ The World Health Organization (WHO) recommends a four-symptom screen (W4SS) for cough, fever, night sweats and/or weight loss prior to ART initiation, followed by rapid molecular TB testing in symptomatic patients.^1^ However, it is logistically challenging to complete a TB evaluation in one day, as the Xpert MTB/RIF assay (Cepheid, Sunnyvale, CA) has at least a two-hour turnaround time, and tests are often conducted in centralized laboratories.^12^ Furthermore, the specificity of the W4SS is low (42% in a recent meta-analysis); consequently, this approach results in additional visits for test results and delays in ART initiation for patients who do not have active TB.

To avoid delays in ART initiation, the current WHO guidelines include a clinical consideration to initiate same-day ART while investigating for TB in patients with presumptive TB (without symptoms of central nervous system involvement).^1^ However, data on the safety of initiating ART in the presence of undiagnosed TB are limited.^13^ A clinical trial is currently underway in patients who report at least one symptom on the W4SS at HIV diagnosis in Lesotho and Malawi to compare outcomes with a strategy of waiting for TB test results vs. immediately initiating ART.^14^

An alternative approach would be to conduct a rapid point-of-care test for symptomatic patients at HIV diagnosis to further improve TB risk profiling, so that TB testing can be prioritized for high-risk patients, and immediate ART can be initiated in low-risk patients. Rapid point-of-care C-reactive protein (CRP) tests could be particularly well suited for this indication, because they are inexpensive ($2 USD/test), generate results in 3 minutes, and can be performed with a fingerstick specimen.^15^ Multiple studies have reported that CRP has superior diagnostic characteristics for the detection of TB at the time of HIV diagnosis, compared with the W4SS alone.^16–19^ The WHO conditionally recommends the use of CRP with a cut-off of 5mg/L to screen for active TB in PLWH, with the caveat that more data are needed.^1^ However, CRP testing is not widely available in HIV clinics, and is not included in decision-making algorithms about the timing of ART initiation. Therefore, we conducted a study to evaluate the association between CRP levels and active TB among symptomatic patients at HIV diagnosis at the Haitian Group for the Study of Kaposi’s Sarcoma and Opportunistic Infections (GHESKIO) in Port-au-Prince, Haiti.

## METHODS

This study included data from a previously published trial, in which persons with non-meningitic TB symptoms at HIV diagnosis were randomized to receive same-day treatment (same-day TB testing with same-day TB treatment if TB diagnosed; same-day ART if TB not diagnosed) vs. standard care (initiating TB treatment within 7 days and delaying ART to Day 7 if TB not diagnosed).^11^ Patients were eligible for study inclusion if they were infected with HIV-1, ≥18 years of age, non-pregnant, ART-naïve, and reported cough, fever, and/or night sweats of any duration, and/or weight loss which was confirmed by the study physician.

At HIV diagnosis, complete blood count (Abbott, Abbott Park, Illinois), CD4 count (FACSCount, Becton Dickinson, Franklin Lakes, NJ), digital chest radiograph, Xpert Ultra (Cepheid, Sunnyvale, CA) and liquid mycobacterial culture (Mycobacteria Growth Indicator Tube [MGIT], BACTEC, Becton Dickinson) were conducted for all participants; Xpert Ultra and mycobacterial culture were performed on both spot and early morning specimens. Sera were stored for CRP testing; specimens were processed within 4 hours of collection, biobanked at −80°C, and shipped to Weill Cornell Medical College for testing (CRP Quantikine ELISA Kit, R&D Systems, Minneapolis, MN). The manufacturer’s protocol was followed with the following specifications. Serum samples were thawed and serially diluted 1:100 followed by 1:5 to make a final dilution of 1:500 for each sample. The 1:100 dilution was saved for subsequent tests if samples needed to be repeated. Absorbance was measured at 450 nm with 540 nm correction. A standard curve (minimum: 3.9 mg/L, maximum: 25 mg/L) and three positive controls with varying concentration were included in each assay performed. Samples that had a CRP reading above 25 mg/L were re-tested at a 1:2,500 or 1:10,000 dilution for each sample.

CRP testing was conducted retrospectively, so results were not available to guide decision making. All participants in both groups received initial TB test results prior to ART or TB treatment initiation; participants in the same-day treatment group received same-day TB test results, which is not standard of care at GHESKIO.

This study was approved by the institutional review boards at GHESKIO, Mass General Brigham, and Weill Cornell Medical College. Written informed consent was obtained from all participants.

### Statistical Analysis

Demographic, clinical and laboratory information were extracted from the GHESKIO electronic medical record. Baseline characteristics were summarized by tuberculosis status using medians and interquartile ranges (IQR) for continuous variables and counts and percentages for categorical variables. Age, body mass index (BMI), hemoglobin, and CD4 count were continuous variables. Sex, income, education, TB symptoms, Xpert Ultra and culture results, and radiographic abnormalities were binary or categorical variables. Patients were classified as having bacteriologically-confirmed TB if they had a positive Xpert Ultra test and/or mycobacterial culture. Patients were classified as having empirically diagnosed TB if they had symptoms and chest radiograph which were consistent with TB, without bacteriologic confirmation.

We calculated sensitivity, specificity, positive and negative predictive values (PPV, NPV), and positive and negative likelihood ratios of varying CRP thresholds (≥1.0, ≥2.0, ≥5.0, ≥10.0 mg/L) in the diagnosis of TB in the total cohort of symptomatic patients, and stratified by symptoms (cough, fever, and/or night sweats vs. weight loss only). We also calculated these test characteristics in the diagnosis of bacteriologically-confirmed TB. In addition, we assessed predictors of high CRP (≥3 mg/dL) in the total cohort, and predictors of high (≥3 mg/L) vs. low (<3 mg/L) CRP among patients diagnosed with TB, using generalized linear models.

## RESULTS

Five hundred participants were enrolled in the trial from November 6, 2017 to January 16, 2020; of these 498 (99.6%) had a CRP test result at HIV diagnosis, and were included in the analysis. Median age was 37 years (IQR: 30, 45), 234 (46.8%) were female, and median body mass index (BMI) was 20.7 (IQR 18.7, 22.9). Median hemoglobin was 11.2 g/dL (IQR: 9.9, 12.3) in females and 12.7 g/dL (10.6, 14.3) in males; median CD4 count was 276 cells/mm^3^ (IQR: 128, 426), and median CRP was 3.9 mg/L (IQR: 1.0, 18.7) (see **Table 1**). A total of 260 participants (52.2%) reported cough, fever, and/or night sweats (+/-weight loss) and 238 (47.8%) reported isolated weight loss.

**Table 1.** Baseline Characteristics by TB Status.

| **Variable** | **Total Cohort**<br>**(n = 498)** | **TB Cohort**<br>**(n= 87)** | **Non-TB Cohort**<br>**(n= 411)** | **p-value** |
| --- | --- | --- | --- | --- |
| Female – no. (%) | 234 (47.0) | 33 (37.9) | 201 (48.9) | 0.081 |
| Age (years) – median (IQR) | 37 (30, 45) | 36 (31, 43) | 37 (30, 46) | 0.531 |
| Cough, fever, and/or night sweats – no. (%) | 260 (52.2) | 78 (89.7) | 182 (44.3) | <0.001* |
| Body mass index – median (IQR) | 20.7 (18.7, 22.9) | 18.9 (17.7, 20.6) | 21.1 (19.2, 23.3) | <0.001* |
| CD4 count category – no. (%) |  |  |  | 0.028 |
| <100 cells/mm<sup>3</sup> | 100 (20.3) | 25 (29.4) | 75 (18.4) |  |
| 100 to 349 cells/mm<sup>3</sup> | 224 (45.4) | 39 (45.9) | 185 (45.3) |  |
| 350 to 499 cells/mm<sup>3</sup> | 84 (17.0) | 14 (16.5) | 70 (17.2) |  |
| ≥500 cells/mm<sup>3</sup> | 85 (17.2) | 7 (8.2) | 78 (19.1) |  |
| CD4 count – median (IQR) | 276 (128, 426) | 170 (86, 342) | 288 (140, 444) | < 0.001* |
| Hemoglobin (g/dL) – median (IQR) | 11.9 (10.1, 13.4) | 9.4 (8.3, 11.0) | 12.2 (10.7, 13.7) | < 0.001* |
| C-Reactive Protein (mg/L) – no. (%) |  |  |  |  |
| ≥1.0 | 371 (74.5) | 83 (95.4) | 288 (70.1) | <0.001* |
| ≥2.0 | 308 (61.8) | 76 (87.4) | 232 (56.4) | <0.001* |
| ≥3.0 | 267 (53.6) | 75 (86.2) | 192 (46.7) | <0.001* |
| ≥5.0 | 226 (45.4) | 66 (75.9) | 160 (38.9) | <0.001* |
| ≥10.0 | 175 (35.1) | 62 (71.3) | 113 (27.5) | <0.001* |
| C-Reactive Protein (mg/L – median (IQR)) | 3.9 (1.0, 18.7) | 33.0 (5.1, 85.5) | 2.6 (0.8, 11.7) | <0.001* |

At enrollment, 87 (17.5%) patients were diagnosed with TB; 67 (13.5%) had bacteriologically-confirmed TB and 20 (4.0%) had empirically-diagnosed TB. Patients diagnosed with baseline TB were more likely to report cough, fever, and/or night sweats (vs. isolated weight loss), and had lower BMI, hemoglobin, and CD4 cell counts, compared to those not diagnosed with TB (see **Table 1**). Median CRP was 33.0 mg/L (IQR: 5.1, 85.5) in those diagnosed with TB, and 2.6 mg/L (IQR: 0.8, 11.7) in those without TB. Any radiographic abnormality consistent with pulmonary TB was detected in 45 (51.7%) of participants diagnosed with TB, and 27 (6.6%) of those without TB.

A total of 267 (53.6%) patients had baseline CRP levels of ≥3.0 mg/L. In multivariable analysis, elevated CRP levels (≥3.0 mg/L) were significantly associated with cough/fever/night sweats vs. isolated weight loss (adjusted odds ratio [aOR] 2.01; 95% confidence interval [CI]: 1.33, 3.07), lower CD4 count (aOR: 0.90 per 50 cells/mm^3^; 95% CI: 0.85, 0.94), lower hemoglobin (aOR: 0.98 per g/dL; 95% CI: 0.97, 1.00) and TB versus no TB diagnosis (aOR:3.50, 95% CI: 1.68, 7.89) (see **Table 2**).

**Table 2.** Predictors of Elevated C-Reactive Protein (. ≥**3.0 mg/dL)**

| Variable | Univariable Analysis |  | Multivariable Analysis |  |
| --- | --- | --- | --- | --- |
|  | Odds Ratio (95% CI) | p-value | Adjusted Odds Ratio (95% CI) | p-value |
| Female Sex | 0.89 (0.63, 1.27) | 0.534 | 0.73 (0.46, 1.15) | 0.176 |
| Age (per decade) | 1.15 (0.96, 1.38) | 0.119 | 1.18 (0.96, 1.45) | 0.114 |
| Cough, fever, and/or night sweats (vs. isolated weight loss) | 2.75 (1.91, 3.96) | <0.001* | 2.01 (1.33, 3.07) | 0.001* |
| Body mass index (per unit) | 0.95 (0.90, 0.99) | 0.024* | 1.04 (0.98, 1.11) | 0.186 |
| CD4 count (per 50 cells/mm<sup>3</sup>) | 0.89 (0.85, 0.92) | <0.001* | 0.90 (0.85, 0.94) | <0.001* |
| Hemoglobin (per g/dL) | 0.97 (0.97, 0.98) | <0.001* | 0.98 (0.97, 1.00) | 0.005* |
| Diagnosed with Tuberculosis | 6.00 (3.11, 12.76) | <0.001* | 3.50 (1.68, 7.89) | 0.001* |

Among patients with TB, we conducted univariable analyses to compare those with lower CRP (<3 mg/L) versus higher CRP (≥3 mg/L). There was no significant difference in any of the variables assessed, including sex, age, symptoms, BMI, CD4 count, hemoglobin, sputum results, or radiographic findings (see **Table 3**).

**Table 3.** Baseline Demographic and Clinical Characteristics among Patients with Tuberculosis, by C-Reactive Protein Level.

| **Variable** | **All Patients with TB (n=87)** | **CRP $\geq 3$ (n=75)** | **CRP $< 3$ (n=12)** | **p-value** |
| --- | --- | --- | --- | --- |
| Female – no. (%) | 33 (37.9) | 28 (37.3) | 5 (41.7) | 0.76 |
| Age at enrollment (years) – median (IQR) | 36 (31, 43) | 36 (31, 42) | 39 (33, 45) | 0.662 |
| Isolated weight loss (no cough, fever, or night sweats) – no (%) | 9 (10.3) | 7 (9.3) | 2 (16.7) | 0.605 |
| Body mass index – median (IQR) | 18.9 (17.7, 20.6) | 18.8 (17.7, 20.7) | 19.4 (17.7, 20.1) | 0.849 |
| CD4 count – median (IQR) | 170 (86, 342) | 177 (88, 317) | 122.5 (70.8, 382.3) | 0.686 |
| CD4 count category – no. (%) |  |  |  | 0.246 |
| <100 cells/mm<sup>3</sup> | 25 (29.4) | 20 (27.4) | 5 (41.7) |  |
| 100 to 349 cells/mm<sup>3</sup> | 39 (45.9) | 36 (49.3) | 3 (25) |  |
| ≥350 cells/mm<sup>3</sup> | 21 (24.7) | 17 (23.2) | 4 (33.4) |  |
| Hemoglobin (g/dL) – median (IQR) | 9.4 (8.3, 11.0) | 9.4 (8.2, 10.9) | 9.1 (8.8., 11.7) | 0.483 |
| Xpert Ultra positive (2 samples) | 37 (42.5) | 33 (44.0) | 4 (33.3) | 0.704 |
| Xpert Ultra positive (1 sample) | 55 (63.2) | 48 (64.0) | 7 (58.3) | 0.753 |
| Xpert Ultra and/or culture positive | 67 (77.0) | 57 (76.0) | 10 (83.3) | 0.725 |
| Any radiographic abnormality consistent with TB | 45 (51.7) | 40 (53.3) | 5 (41.7) | 0.66 |

We assessed the impact of a range of CRP thresholds on the PPV and NPV of baseline TB in this cohort of symptomatic patients (see **Table 4**). As the CRP threshold increased from ≥1 mg/L, to ≥3 mg/L, ≥5 mg/L and ≥10 mg/L, the PPV for TB in the total cohort increased from 22.4% to 28.1%, 29.2%, and 35.4%, respectively, and the NPV decreased from 96.9% to 94.8%, 92.3%, and 92.3%. **Supplementary Table 1** includes these results in patients with bacteriologically-confirmed TB.

**Table 4.** Utility of C-Reactive Protein Thresholds for Ruling In or Ruling Out Tuberculosis.

| **CRP Threshold** | **Proportion of Total Study Population (%)** | **Sensitivity (95% CI)** | **Specificity (95% CI)** | **PPV (95% CI)** | **NPV (95% CI)** | **Positive LR (95% CI)** | **Negative LR (95% CI)** |
| --- | --- | --- | --- | --- | --- | --- | --- |
| ***Total Cohort (Patients with Cough, Fever, Night Sweats, and/or Weight Loss)*** |  |  |  |  |  |  |  |
| ≥1.0 | 74.5 | 95.4 (88.6-98.7) | 29.9 (25.5-34.6) | 22.4 (18.2-27.0) | 96.9 (92.1-99.1) | 1.4 (1.3-1.5) | 0.2 (0.1-0.4) |
| ≥2.0 | 61.8 | 87.4 (78.5-93.5) | 43.6 (38.7-48.5) | 24.7 (20-29.9) | 94.2 (89.9-97.1) | 1.5 (1.4-1.7) | 0.3 (0.2-0.5) |
| ≥3.0 | 53.6 | 86.2 (77.1-92.7) | 53.3 (48.3-58.2) | 28.1 (22.8-33.9) | 94.8 (91.1-97.3) | 1.8 (1.6-2.1) | 0.3 (0.2-0.4) |
| ≥5.0 | 45.4 | 75.9 (65.5-84.4) | 61.1 (56.2-65.8) | 29.2 (23.4-35.6) | 92.3 (88.4-95.2) | 1.9 (1.6-2.3) | 0.4 (0.3-0.6) |
| ≥10.0 | 35.1 | 71.3 (60.6-80.5) | 72.5 (67.9-76.8) | 35.4 (28.4-43.0) | 92.3 (88.8-94.9) | 2.6 (2.1-3.2) | 0.4 (0.3-0.6) |
| ***Patients with Cough, Fever, and/or Night Sweats +/- Weight Loss (Excludes Patients with Isolated Weight Loss)*** |  |  |  |  |  |  |  |
| ≥1.0 | 81.9 | 94.9 (87.4-98.6) | 23.6 (17.7-30.5) | 34.7 (28.4-41.5) | 91.5 (79.6-97.6) | 1.2 (1.1-1.4) | 0.2 (0.1-0.6) |
| ≥2.0 | 71.9 | 87.2 (77.7-93.7) | 34.6 (27.7-42) | 36.4 (29.5-43.7) | 86.3 (76.2-93.2) | 1.3 (1.2-1.5) | 0.4 (0.2-0.7) |
| ≥3.0 | 65.4 | 87.2 (77.7-93.7) | 44.0(36.6-51.5) | 40.0 (32.6-47.8) | 88.9 (80.5-94.5) | 1.6 (1.3-1.8) | 0.3 (0.2-0.5) |
| ≥5.0 | 58.8 | 80.8 (70.3-88.8) | 50.5 (43.1-58) | 41.2 (33.3-49.4) | 86.0 (77.9-91.9) | 1.6 (1.4-2.0) | 0.4 (0.2-0.6) |
| ≥10.0 | 49.6 | 75.6 (64.6-84.7) | 61.5 (54.1-68.6) | 45.7 (36.9-54.7) | 85.5 (78.3-91.0) | 2.0 (1.6-2.5) | 0.4 (0.3-0.6) |
| ***Patients with Isolated Weight Loss (Excludes Patients with Cough, Fever, or Night Sweats)*** |  |  |  |  |  |  |  |
| ≥1.0 | 66.4 | 100 (66.4-100) | 34.9 (28.8-41.5) | 5.7 (2.6-10.5) | 100.0 (95.5-100.0) | 1.5 (1.4-1.7) | 0 |
| ≥2.0 | 50.8 | 88.9 (51.8-99.7) | 50.7 (44.0-57.3) | 6.6 (2.9-12.6) | 99.1 (95.3-100.0) | 1.8 (1.4-2.3) | 0.2 (0.0-1.4) |
| ≥3.0 | 40.8 | 77.8 (40.0-97.2) | 60.7 (54.0-67.1) | 7.2 (3.0-14.3) | 98.6 (95.0-99.8) | 2 (1.3-2.9) | 0.4 (0.1-1.2) |
| ≥5.0 | 30.7 | 33.3 (7.5-70.1) | 69.4 (63.0-75.3) | 4.1 (0.9-11.5) | 96.4 (92.3-98.7) | 1.1 (0.4-2.8) | 1 (0.6-1.5) |
| ≥10.0 | 19.3 | 33.3 (7.5-70.1) | 81.2 (75.6-86.1) | 6.5 (1.4-17.9) | 96.9 (93.3-98.8) | 1.8 (0.7-4.6) | 0.8 (0.5-1.3) |

Abbreviations: LR: Likelihood ration; NPV: negative predictive value; PPV positive predictive value

Among the 260 participants with cough, fever, and/or night sweats (+/-weight loss), the prevalence of baseline TB was 30%. The PPV for TB increased from 34.7% to 40.0%, 41.2%, and 45.7%, as the CRP threshold increased from ≥1 mg/L, to ≥3 mg/L, ≥5 mg/L and ≥10 mg/L, respectively, and the NPV decreased from 91.5% to 88.9%, 86.0%, and 85.5%.

Among the 238 participants with isolated weight loss (i.e., excluding those with cough, fever, and/or night sweats), the prevalence of TB was 3.8%. The PPV was only 6.5% at CRP threshold of ≥10 mg/L. The NPV for TB decreased from 100% at a CRP threshold of ≥1 mg/L, to 98.6%, 96.4%, and 96.9% at thresholds of ≥3 mg/L, ≥5 mg/L and ≥10 mg/L, respectively.

CRP testing was conducted retrospectively, so results were not available at enrollment in the parent trial; in accordance with the study protocol, all 498 patients had TB testing prior to ART initiation. If we had implemented the WHO-recommended strategy of ART initiation prior to completion of TB testing, then 87 (17.5%) patients would have started ART with untreated TB. If we had included CRP concentrations in our same-day ART eligibility algorithm, then with CRP thresholds of <1, <2, <3, < 5, and <10 mg/L, a total of 127 (25.5%), 190 (38.2%), 231 (46.4%), 272 (54.6%), and 323 (64.9%) patients would have been eligible for same-day ART (see **Figure 1**). A total of 4 (0.8%), 11 (2.2%), 12 (2.4%), 21 (4.2%), and 25 (5.0%) patients, respectively, would have had untreated active TB at ART initiation (active TB in spite of having CRP levels below the cut-off).

**Figure 1.**
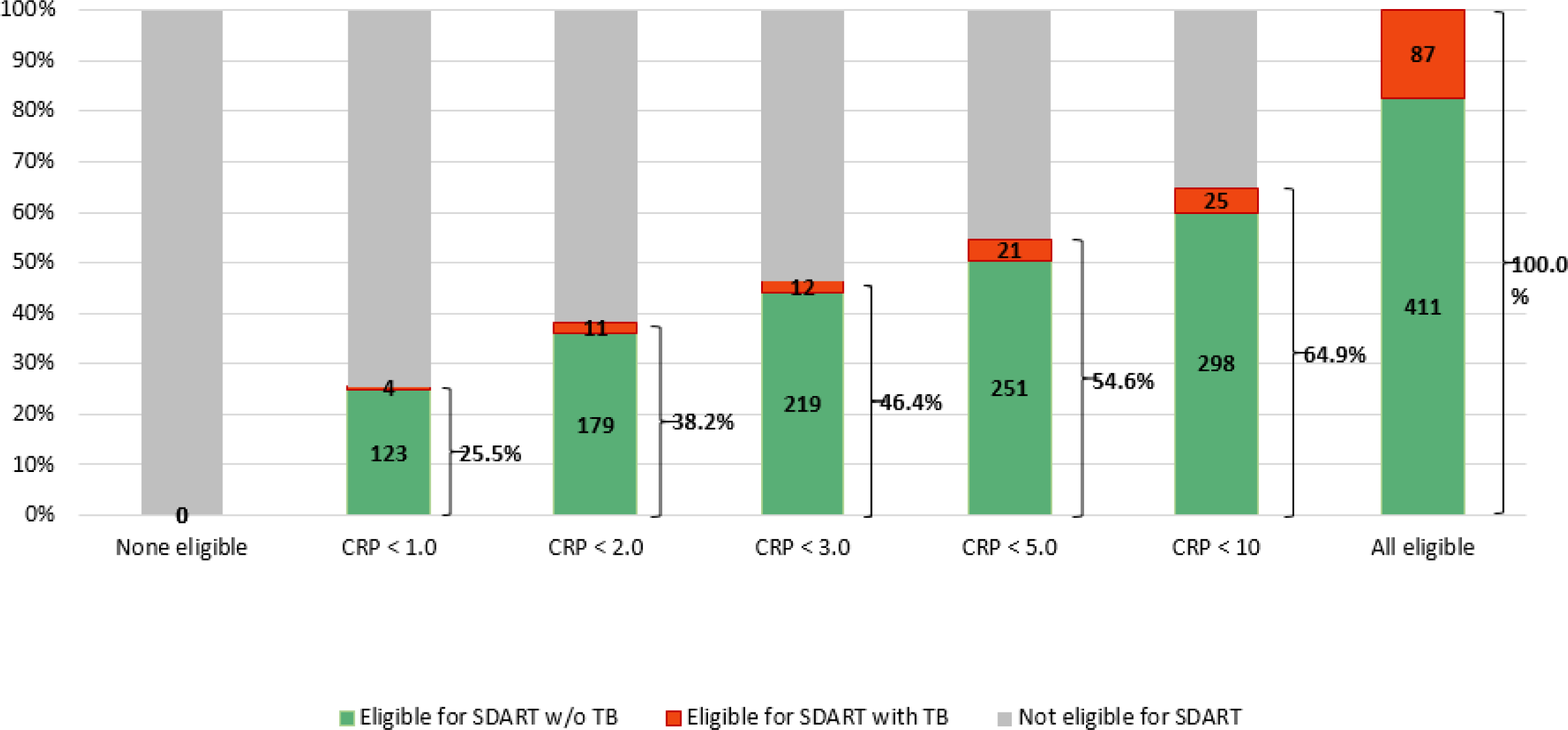
Impact of CRP-Based ART Eligibility Thresholds on Same-Day ART Qualification and Undiagnosed Tuberculosis (n=498)

Among the 260 patients who reported cough, fever, and/or night sweats (+/-weight loss), if same-day ART had been initiated while TB testing was underway, then 78 (30.0%) would have started ART with untreated TB. If CRP thresholds of <1, <2, <3, <5, and <10 mg/L had been used as eligibility thresholds, then 47 (17.5%), 73 (28.1%), 90 (34.6%), 107 (41.2%), and 131 (50.4%) patients, respectively would have been eligible for same-day ART (see **Figure 2A**). A total of 4 (1.5%), 10 (3.8%), 10 (3.8%), 15 (5.8%), and 19 (7.3%) patients, respectively, would have had untreated TB at ART initiation.

**Figure 2A.**
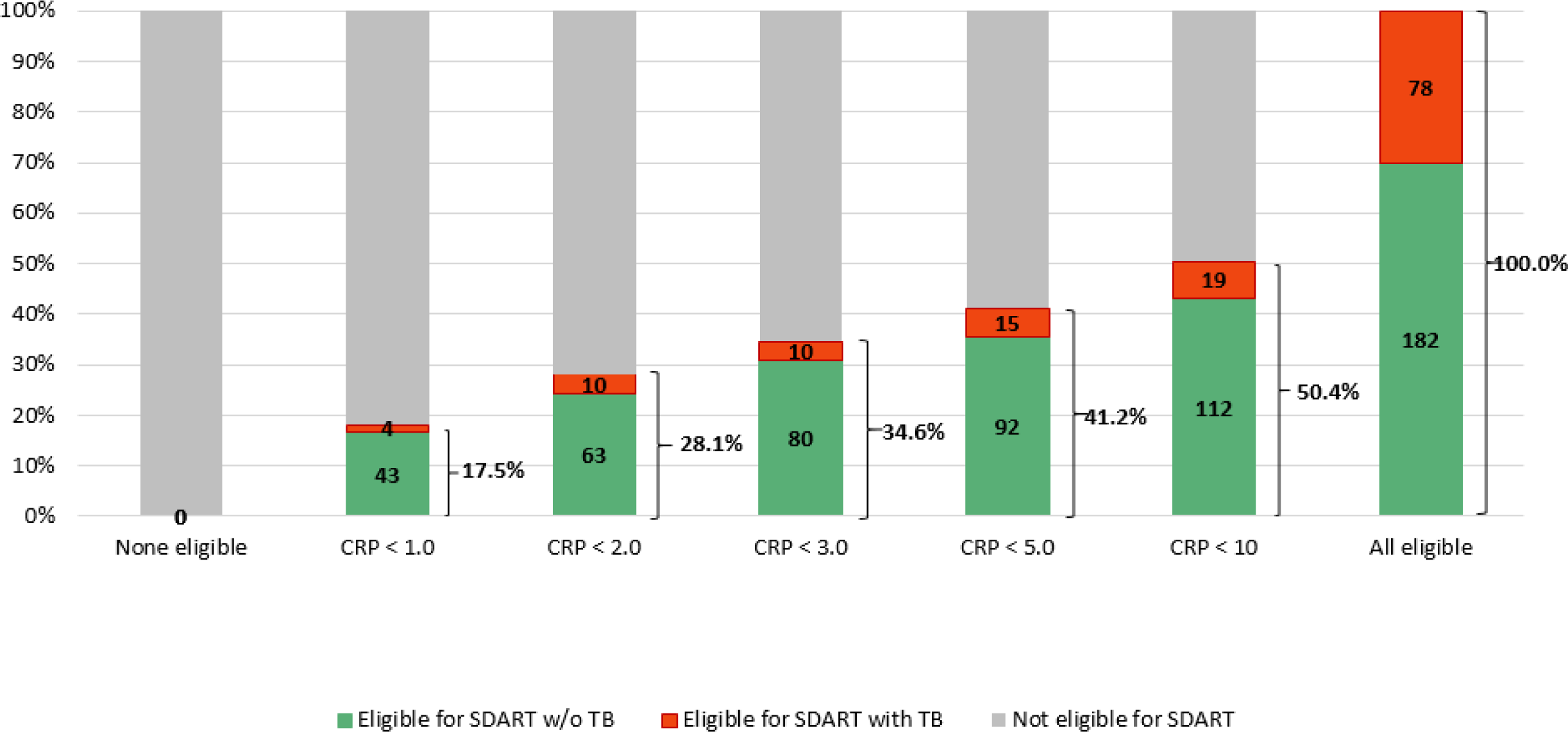
Impact of CRP-Based Eligibility Criteria on Same-Day ART Eligibility and Untreated TB in Patients with Cough, Fever, and/or Night Sweats at HIV Diagnosis (n=260)

Among the 238 patients who reported isolated weight loss, 9 (3.8%) were diagnosed with baseline TB. If CRP thresholds of <1, <2, <3, <5, and <10 mg/L had been used as same-day ART eligibility thresholds, then 80 (33.6%), 117 (49.2%), 141 (59.2%), 165 (69.3%), and 192 (80.7%) patients, respectively would have been eligible for same-day ART (see **Figure 2B**). A total of 0 (0.0%), 1 (0.4%), 2 (0.8%), 6 (2.5%), and 6 (2.5%) patients, respectively, would have had untreated TB at ART initiation.

**Figure 2B.**
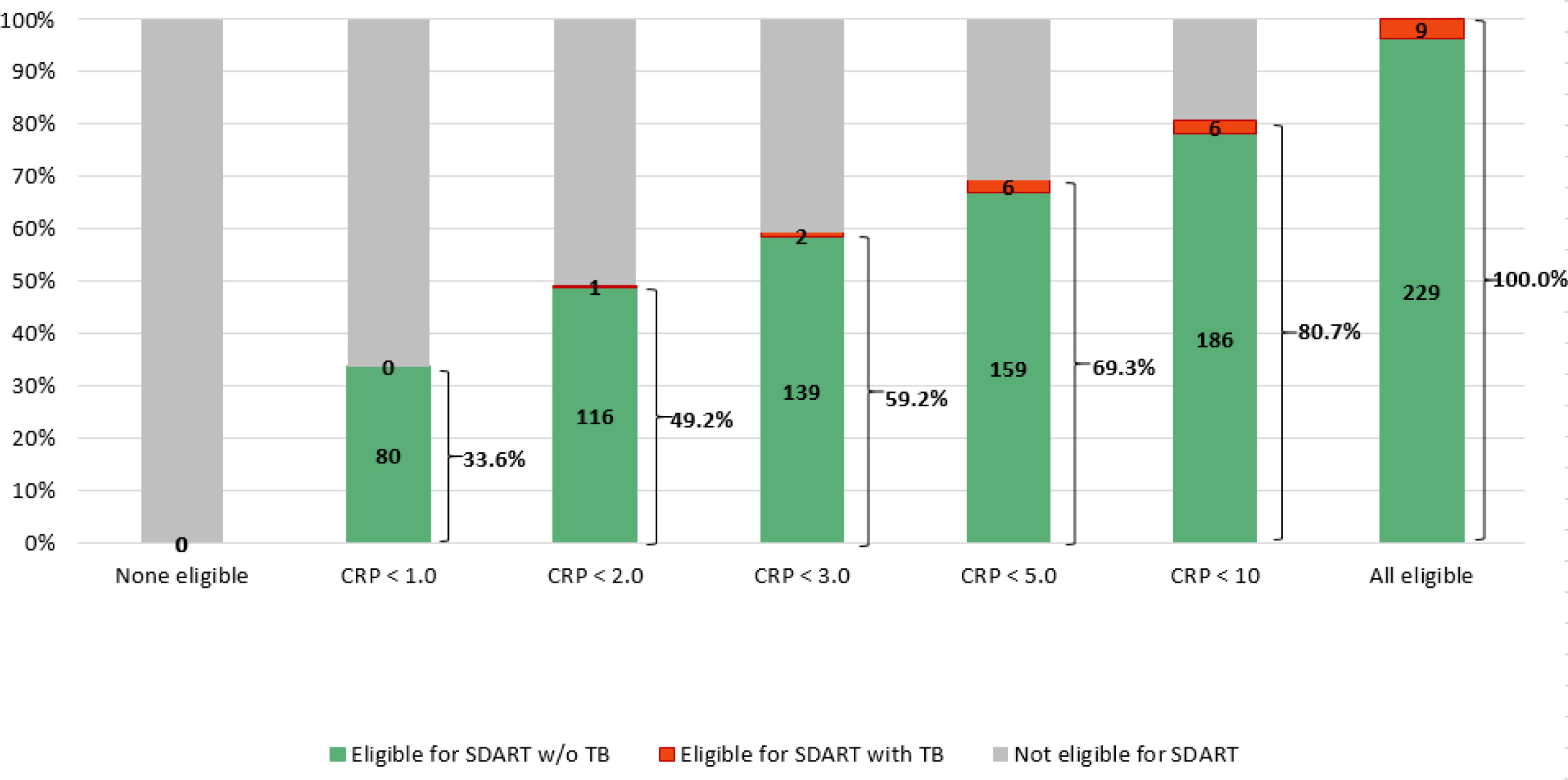
Impact of CRP-Based Eligibility Criteria on Same-Day ART Eligibility and Untreated TB in Patients with Isolated Weight Loss at HIV Diagnosi s (n=238)

## DISCUSSION

We found that CRP concentrations can be used to stratify symptomatic patients into high or low risk of TB at HIV diagnosis, facilitating decision making about same-day ART initiation. The WHO currently recommends the W4SS screen at HIV diagnosis, with two guideline-approved algorithms for symptomatic patients – either same-day ART while awaiting TB test results, or deferral of ART until completion of TB testing.^1^ However, these approaches either result in ART initiation in patients with undiagnosed TB, or unnecessary delays in ART for those without TB. In our cohort, the prevalence of baseline TB was 17.5%, and the PPV of CRP was 22.4%, 28.1%, 29.2%, and 35.4% at thresholds ≥1 mg/L, ≥3 mg/L, ≥5 mg/L and ≥10 mg/L, respectively. With the use of CRP-based eligibility criteria, one-quarter to two-thirds of symptomatic patients could be eligible for same-day ART, depending on the CRP threshold used, with a 3-fold to 20-fold reduction in the proportion of patients with undiagnosed TB at ART initiation, compared with the WHO strategy of same-day ART prior to completion of TB testing.

We also found that baseline TB prevalence varied by W4SS symptom category. About half of the total cohort reported cough, fever, and/or nights sweats (+/-weight loss), and nearly half reported isolated weight loss. The prevalence of TB in those with cough, fever, and/or night sweats was almost 10-fold higher than that of the patients who reported isolated weight loss (30.0% vs. 3.8%). Among patients with cough, fever, and/or night sweats, the PPV of CRP was 40.0%, 41.2%, and 45.7% at thresholds ≥3 mg/L, ≥5 mg/L and ≥10 mg/L, respectively. If CRP values under these thresholds were used to define same-day ART eligibility, then one-third to one-half of this subgroup would be eligible for same-day ART, with a 4-fold to 8-fold reduction in the proportion of patients with undiagnosed TB at ART initiation, compared with the strategy of same-day ART testing prior to completion of TB testing.

In contrast, among patients with isolated weight loss, the PPV is low at all CRP thresholds, but the NPV is high; with CRP thresholds of <1, <2, or <3, up to about 60% of patients would be eligible for same-day ART initiation, with <1% of patients having baseline TB and CRP concentrations below these cut-offs. A study conducted among ART-naïve patients in South Africa also reported that the NPV of CRP <1 mg/L was 100%.^20^ These data suggest that clinicians can confidently commence ART in patients with a new diagnosis of HIV with isolated weight loss as a symptom and low CRP. In fact, from our study, less than 1% of patients with this clinical picture and a CRP <3 mg/L were found to have TB.

Multiple studies of ART-naïve patients in African cohorts that included mycobacterial culture have reported a TB prevalence similar to ours.^21–25^ Most studies that reported 4WSS results combined all four symptoms.^26^ Among those that reported individual symptoms, other studies have also reported a lower TB prevalence among patients reporting isolated weight loss, compared with cough, fever, and/or night sweats. In a clinical trial cohort of newly diagnosed patients from South Africa and Kenya, the PPV value of cough and fever was over 30% versus about 5% for isolated weight loss.^27^ A cohort study from Ethiopia found that cough was the most predictive of active TB.^25^

We are not aware of any HIV program which uses CRP in decision making about the timing of ART initiation. Multiple studies have demonstrated CRP has similar sensitivity but greater specificity than the W4SS, but these studies were conducted to assess alternative strategies for more efficient use of TB diagnostic testing.^16–20^ Current WHO guidelines provide a conditional recommendation (low-certainty evidence for test accuracy) for CRP with a cut-off of >5 mg/L to screen for TB disease in PLWH.^1^ However, CRP is not included in the algorithms for same-day ART initiation.

The sensitivity and specificity of the W4SS are about 82% and 42%, respectively.^19,26^ In a systematic review and individual participant data meta-analysis commissioned by the WHO, the investigators found that among outpatients not on ART, the sensitivities of CRP (≥10 mg/L) and a sequential strategy of W4SS followed by CRP (≥5 mg/L) were similar to W4SS alone, but the specificities were higher.^19^ A study from Uganda found that point-of-care CRP had 89% sensitivity and 72% specificity for culture-confirmed TB.^17^ A study from South Africa reported that the sensitivity and specificity of CRP >5 for culture-positive TB were 91% and 59%, respectively.^16^ Among our cohort of patients with positive W4SS symptom screens, the sensitivity of CRP was higher at threshold of ≥3 mg/L versus ≥5 mg/L (86% vs. 76%), with a slight decline in specificity (53% vs. 61%).^19^

These findings highlight the potential utility of CRP testing to inform decision making about same-day ART initiation in patients who present with TB symptoms at HIV diagnosis. CRP is available as a simple, rapid, low-cost (<$US 2), point-of-care test, which is available from a variety of manufacturers.^15^ Further study is necessary to define optimal CRP thresholds, which may potentially vary by number and type of symptoms. Moreover, CRP would not replace diagnostic testing for TB or other opportunistic infections and non-communicable diseases.

Our study was conducted among non-pregnant participants in a large, urban clinic, which may limit the generalizability of our findings. It is also important to note that we used a more aggressive diagnostic approach for TB than is provided in many high-burden settings. However, in settings without access to mycobacterial culture and chest radiographs, the detection of a high CRP level (>5mg/L or >10 mg/L for example) can alert providers to a potential case of TB or other opportunistic infection, even if sputum smear or Xpert MTB/RIF testing is negative. Among patients with isolated weight loss without respiratory symptoms or fever, the clinician and program can have even greater confidence in commencing ART (and not missing a TB diagnosis) in the presence of a low CRP.

In conclusion, our results demonstrate that CRP testing can be useful in stratifying patients with non-meningitic symptoms into high or low risk of TB at HIV diagnosis, facilitating decision making about same-day ART initiation.

## Data Availability

All data produced in the present study are available upon reasonable request to the authors

## ACKNOWLEDGEMENTS

We thank the patients who participated in this study and the GHESKIO staff who cared for them. We also thank the members of the Data Safety Monitoring Board, the Community Advisory Board, and the ethics committees for their expertise and advice. This study was funded by a grant from the National Institute of Allergy and Infectious Diseases (R01AI131998; primary investigator: SK). KD was supported by NIAID (K23AI131913) and a Doris Duke Charitable Foundation Clinical Scientist Development Award.

All authors were involved in drafting the article, and all approved the final version to be published. Study conception and design: KD, VRR, ND, CR, PS, SP, KFW, PJ, JWP, SPK. Acquisition of data: ND, AA, ED, CR, PS, KL, SP, VR. Analysis and interpretation of data: KD, VRR, ND, HA, YG, JL, MDM, KFW, ALB, PYC, JWP, SPK. There are no conflicts of interest.

---

**Supplemental Table 1.**
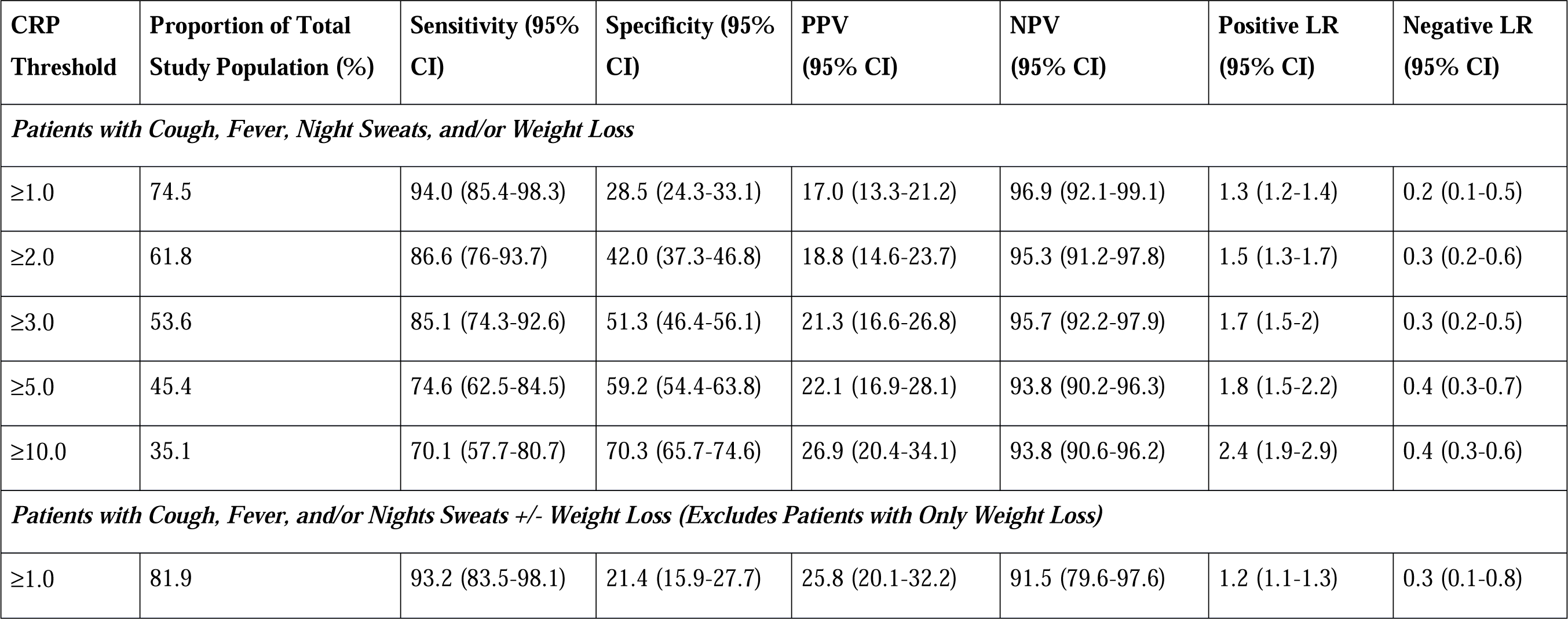
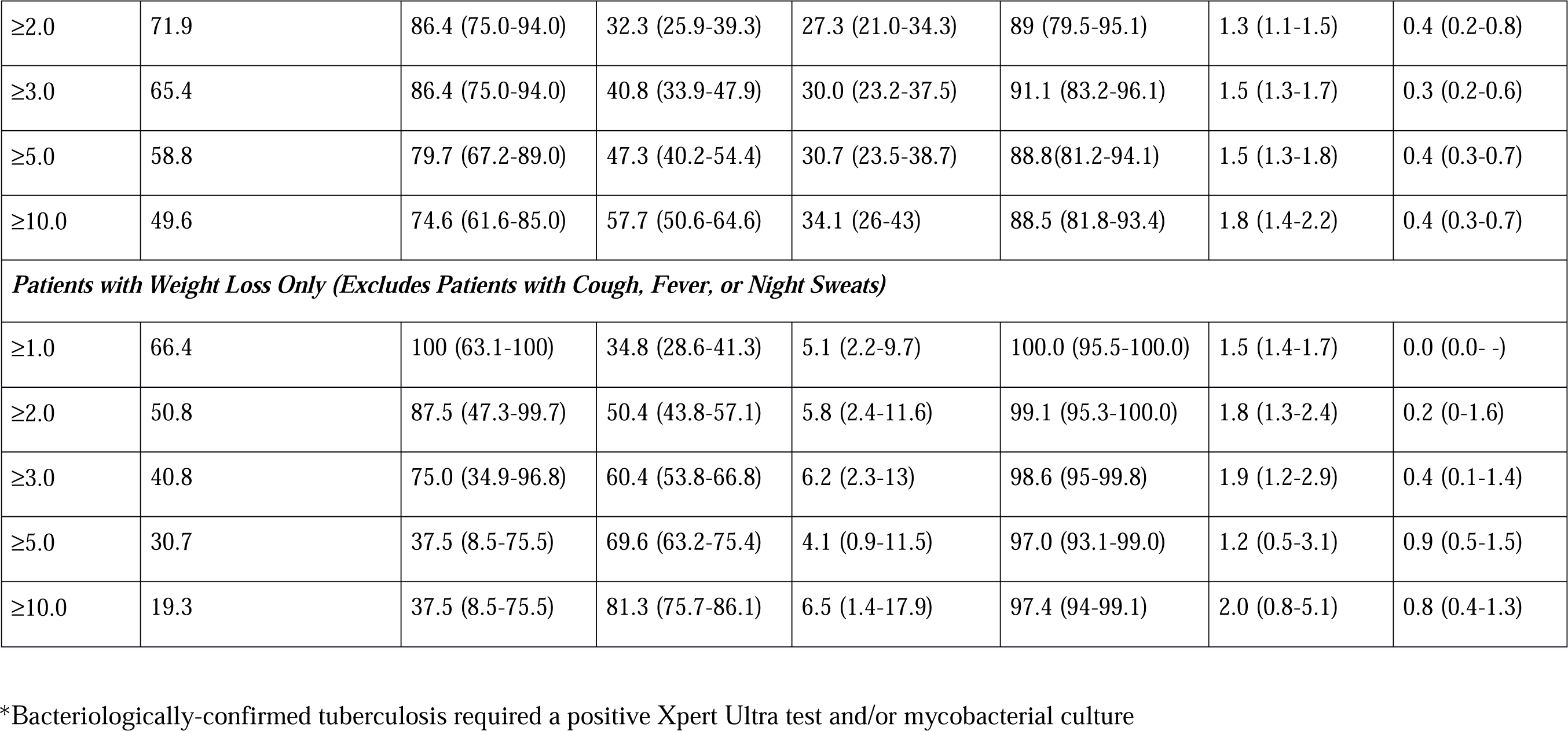
Utility of C-Reactive Protein Thresholds for Ruling In or Ruling Out Bacteriologically-Confirmed Tuberculosis*.

| **CRP Threshold** | **Proportion of Total Study Population (%)** | **Sensitivity (95% CI)** | **Specificity (95% CI)** | **PPV (95% CI)** | **NPV (95% CI)** | **Positive LR (95% CI)** | **Negative LR (95% CI)** |
| --- | --- | --- | --- | --- | --- | --- | --- |
| ***Patients with Cough, Fever, Night Sweats, and/or Weight Loss*** |  |  |  |  |  |  |  |
| ≥1.0 | 74.5 | 94.0 (85.4-98.3) | 28.5 (24.3-33.1) | 17.0 (13.3-21.2) | 96.9 (92.1-99.1) | 1.3 (1.2-1.4) | 0.2 (0.1-0.5) |
| ≥2.0 | 61.8 | 86.6 (76-93.7) | 42.0 (37.3-46.8) | 18.8 (14.6-23.7) | 95.3 (91.2-97.8) | 1.5 (1.3-1.7) | 0.3 (0.2-0.6) |
| ≥3.0 | 53.6 | 85.1 (74.3-92.6) | 51.3 (46.4-56.1) | 21.3 (16.6-26.8) | 95.7 (92.2-97.9) | 1.7 (1.5-2) | 0.3 (0.2-0.5) |
| ≥5.0 | 45.4 | 74.6 (62.5-84.5) | 59.2 (54.4-63.8) | 22.1 (16.9-28.1) | 93.8 (90.2-96.3) | 1.8 (1.5-2.2) | 0.4 (0.3-0.7) |
| ≥10.0 | 35.1 | 70.1 (57.7-80.7) | 70.3 (65.7-74.6) | 26.9 (20.4-34.1) | 93.8 (90.6-96.2) | 2.4 (1.9-2.9) | 0.4 (0.3-0.6) |
| ***Patients with Cough, Fever, and/or Night Sweats +/- Weight Loss (Excludes Patients with Only Weight Loss)*** |  |  |  |  |  |  |  |
| ≥1.0 | 81.9 | 93.2 (83.5-98.1) | 21.4 (15.9-27.7) | 25.8 (20.1-32.2) | 91.5 (79.6-97.6) | 1.2 (1.1-1.3) | 0.3 (0.1-0.8) |
| ≥2.0 | 71.9 | 86.4 (75.0-94.0) | 32.3 (25.9-39.3) | 27.3 (21.0-34.3) | 89 (79.5-95.1) | 1.3 (1.1-1.5) | 0.4 (0.2-0.8) |
| ≥3.0 | 65.4 | 86.4 (75.0-94.0) | 40.8 (33.9-47.9) | 30.0 (23.2-37.5) | 91.1 (83.2-96.1) | 1.5 (1.3-1.7) | 0.3 (0.2-0.6) |
| ≥5.0 | 58.8 | 79.7 (67.2-89.0) | 47.3 (40.2-54.4) | 30.7 (23.5-38.7) | 88.8(81.2-94.1) | 1.5 (1.3-1.8) | 0.4 (0.3-0.7) |
| ≥10.0 | 49.6 | 74.6 (61.6-85.0) | 57.7 (50.6-64.6) | 34.1 (26-43) | 88.5 (81.8-93.4) | 1.8 (1.4-2.2) | 0.4 (0.3-0.7) |
| ***Patients with Weight Loss Only (Excludes Patients with Cough, Fever, or Night Sweats)*** |  |  |  |  |  |  |  |
| ≥1.0 | 66.4 | 100 (63.1-100) | 34.8 (28.6-41.3) | 5.1 (2.2-9.7) | 100.0 (95.5-100.0) | 1.5 (1.4-1.7) | 0.0 (0.0- -) |
| ≥2.0 | 50.8 | 87.5 (47.3-99.7) | 50.4 (43.8-57.1) | 5.8 (2.4-11.6) | 99.1 (95.3-100.0) | 1.8 (1.3-2.4) | 0.2 (0-1.6) |
| ≥3.0 | 40.8 | 75.0 (34.9-96.8) | 60.4 (53.8-66.8) | 6.2 (2.3-13) | 98.6 (95-99.8) | 1.9 (1.2-2.9) | 0.4 (0.1-1.4) |
| ≥5.0 | 30.7 | 37.5 (8.5-75.5) | 69.6 (63.2-75.4) | 4.1 (0.9-11.5) | 97.0 (93.1-99.0) | 1.2 (0.5-3.1) | 0.9 (0.5-1.5) |
| ≥10.0 | 19.3 | 37.5 (8.5-75.5) | 81.3 (75.7-86.1) | 6.5 (1.4-17.9) | 97.4 (94-99.1) | 2.0 (0.8-5.1) | 0.8 (0.4-1.3) |

\*Bacteriologically-confirmed tuberculosis required a positive Xpert Ultra test and/or mycobacterial culture

